# Repeated vaccination with homologous influenza hemagglutinin broadens human antibody responses to unmatched flu viruses

**DOI:** 10.1101/2024.03.27.24303943

**Authors:** Yixiang Deng, Melbourne Tang, Ted M. Ross, Aaron G. Schmidt, Arup K. Chakraborty, Daniel Lingwood

**Affiliations:** Ragon Institute of Mass General, MIT, and Harvard, Cambridge, MA; Department of Physics, Massachusetts Institute of Technology, Cambridge, MA; Center for Vaccines and Immunology, University of Georgia, Athens, GA; Department of Infectious Diseases, University of Georgia, Athens, GA; Department of Microbiology, Harvard Medical School, Boston, MA; Department of Chemical Engineering, Massachusetts Institute of Technology, Cambridge, MA 02139; Department of Chemistry, Massachusetts Institute of Technology, Cambridge, MA 02139; Institute for Medical Engineering and Science, Massachusetts Institute of Technology, Cambridge, MA 02139

## Abstract

The on-going diversification of influenza virus necessicates annual vaccine updating. The vaccine antigen, the viral spike protein hemagglutinin (HA), tends to elicit strain-specific neutralizing activity, predicting that sequential immunization with the same HA strain will boost antibodies with narrow coverage. However, repeated vaccination with homologous SARS-CoV-2 vaccine eventually elicits neutralizing activity against highly unmatched variants, questioning this immunological premise. We evaluated a longitudinal influenza vaccine cohort, where each year the subjects received the same, novel H1N1 2009 pandemic vaccine strain. Repeated vaccination gradually enhanced receptor-blocking antibodies (HAI) to highly unmatched H1N1 strains within individuals with no initial memory recall against these historical viruses. An *in silico* model of affinity maturation in germinal centers integrated with a model of differentiation and expansion of memory cells provides insight into the mechanisms underlying these results and shows how repeated exposure to the same immunogen can broaden the antibody response against diversified targets.

## Introduction

Seasonal influenza vaccines are designed to elicit protective antibody responses against the viral strains predicted to dominate during an upcoming winter season (Comber et al., 2023; Fiore et al., 2009; Jordan et al., 2023; Sandor et al., 2021). The vaccine is typically trivalent or quadrivalent and aims to cover human-infecting influenza A viruses (IAV) and influenza B viruses (IBV) (Demirden et al., 2022; Reed et al., 2012; Soema et al., 2015). This has included an H1N1 vaccine strain for group 1 IAV, an H3N2 vaccine strain for group 2 IAV, and Yamagata and/or Victoria lineages for IBV. Elicitation of antibodies engaging the receptor binding site (RBS) on the influenza spike protein hemagglutinin (HA) to block viral attachment is considered a major source of protection and is routinely measured by the hemagglutination (HA) inhibition (HAI) assay (Cox, 2013; Krammer et al., 2020; Pedersen, 2014; Spackman and Sitaras, 2020). Antibody Fc effector functions also provide orthogonal immuno-protective activities (Boudreau and Alter, 2019; Boudreau et al., 2023; DiLillo et al., 2016).

A concern of seasonal influenza vaccines is the lack of universality, where vaccine coverage can be lowered by antigenic drift of the virus, or even more worrying, antigen shift leading to the emergence of pandemic flu strains (Bedi et al., 2023; Krammer et al., 2018). These limitations are also underscored by the fact that individual HA molecules tend to elicit strain-specific antibody binding or neutralizing activity (Altman et al., 2018; Angeletti and Yewdell, 2018; Bedi *et al*., 2023; Sangesland and Lingwood, 2021). Here, sequential immunization with homologous influenza HA antigens typically serves to boost strain-limited humoral output (Henry et al., 2018; Krammer *et al*., 2018; Krammer and Palese; Sangesland and Lingwood, 2021). Addressing these deficits has been a basis for rationally designed immune-focusing concepts tasked with re-orienting humoral immunity upon immune subdominant sites of conservation on influenza HA (Altman *et al*., 2018; Angeletti and Yewdell, 2018; Caradonna and Schmidt, 2021; Krammer *et al*., 2018; Sangesland and Lingwood, 2021; Wei et al., 2020). These efforts include structure-based reconfiguration and presentation of conserved HA moieties, and sequential immunization with strain variant antigens to further promote expansion of B cell memory against the invariant sites (Amitai et al., 2020; Angeletti et al., 2019; Boyoglu-Barnum et al., 2021; Caradonna et al., 2022; Nachbagauer and Palese, 2018; Sangesland et al., 2019; Yassine et al., 2015). A number of these ‘universal’ vaccine candidates are at various stages of clinical evaluation (Andrews et al., 2023; Nachbagauer et al., 2021; Widge et al., 2023).

Notably however, recent human SARS-CoV-2 vaccine data warrants reconsideration of the basic premise that sequential immunization with homologous antigens elicits strong but strain-limited humoral immunity (Garcia-Beltran et al., 2022; Muecksch et al., 2022; Schmidt et al., 2022). Three sequential vaccinations with the homologous Wuhan-strain glycoprotein spike antigen elicits neutralizing antibody responses against highly unmatched Omicron variants. Broad neutralizing activity via engagement of the SARS-CoV-2 receptor binding site was acquired after the third vaccination, consistent with a diversification of the repertoire of the antibodies elicited (Garcia-Beltran *et al*., 2022; Muecksch *et al*., 2022; Schmidt *et al*., 2022). Both antigen presentation dynamics and epitope masking activities within B cell germinal centers (GCs) appear to play key roles in the emergence of this broadened antibody response (Yang et al., 2023).

In the present study, we evaluated whether diversification of antibody binding/neutralization breath via sequential immunization with homologous antigen is a general human vaccine principle. Accordingly, we evaluated an influenza vaccine cohort of individuals sampled longitudinally over four years (2013-2016) (Boudreau *et al*., 2023; Nunez et al., 2017). HAI was measured before and after vaccination in each year, using a virus panel composed of diverse influenza A and B viruses spanning > 100 years of evolution (Nunez *et al*., 2017). Importantly, this vaccine cohort closely followed the 2009-2010 H1N1 pandemic and included 4 years of repeat exposure to ‘non-imprinted’ /pandemic A/California/7/2009 (pHA) as the sole H1N1 vaccine strain. Annual vaccination boosted HAI to vaccine matched virus but also, to highly divergent H1N1 viruses, despite the strong lack of relatedness. Importantly this broadening did not occur via initial memory recall, but rather intensified gradually, over the four year vaccination period within individuals that were devoid of initial back-boosting against historical H1N1 viruses. To define a mechanistic framework for this effect, we extended a previous *in silico* model that accounts for B cell affinity maturation within GCs and associated memory B cell differentiation and expansion outside germinal centers (Yang *et al*., 2023). Using this approach, we describe mechanisms that underlie the broadening of antibody coverage. We find that the broadening of the response is determined by the interplay between enhanced antigen presentation and epitope masking in germinal centers after booster shots, germline B cell affinities for different HA epitopes, and the level of conservation of these epitopes in the vaccinating strain with those on different historical vaiants. In these contexts, the capacity to eventually elicit broadly reactive antibody responses using a single influenza vaccine strain is discussed.

## Results

### The RBS Patch of the 2009 H1N1 pandemic virus is strongly divergent from prior influenza strains

We began by applying a structure based approach to define amino acid variation within the RBS (the epitope patch responsible for conferring HAI) amongst diverse IAV (H3N2 and H1N1) and IBV, spanning over 100 years of evolution (Figure 1, Table S1, Data S1). We assessed amino acid relatedness of the residues comprising the entire HA ectodomain (Figure 1A,B), and then the RBS patch, as defined by the structures of four human broadly neutralizing RBS-directed antibodies (bnAbs), each in co-complex with HA (Schmidt et al., 2015) (Figure 1C,D). The paratopes of these bnAbs structurally mimic sialic acid, the primary receptor for influenza virus (Schmidt *et al*., 2015). In each case, the epitope footprint consists of the core viral amino acid residues responsible for binding sialyl oligosaccharide, along with a surrounding ‘ring’ of contact positions that are differentially engaged by the four bnAbs (Figure 1C). We defined the RBS patch as the sialic acid binding residues + the cumulative ‘ring’ of contact positions defined by these antibodies (Figure 1C). Amino acid relatedness values within the HA ectodomain and the RBS patch were then represented as heat maps for the influenza A and B viruses (Figure 1B,D). Within H1N1 viruses, the 2009 pandemic strain (pHA) stands out, along with A/New Jersey/1976, as strongly divergent, particularly within the RBS patch (Figure 1B,D). This is consistent with previous reports on the structure of the RBS (Cheung et al., 2020; Hong et al., 2013; Xu et al., 2010) and the fact that both 2009 pandemic virus and the 1976 outbreak in Fort Dix, New Jersey, originated from swine lineages of H1N1 (Garten et al., 2009; Gaydos et al., 2006; Mena et al., 2016; Sencer, 2011; Smith et al., 2009; Zimmer and Burke, 2009).

**Figure 1.**
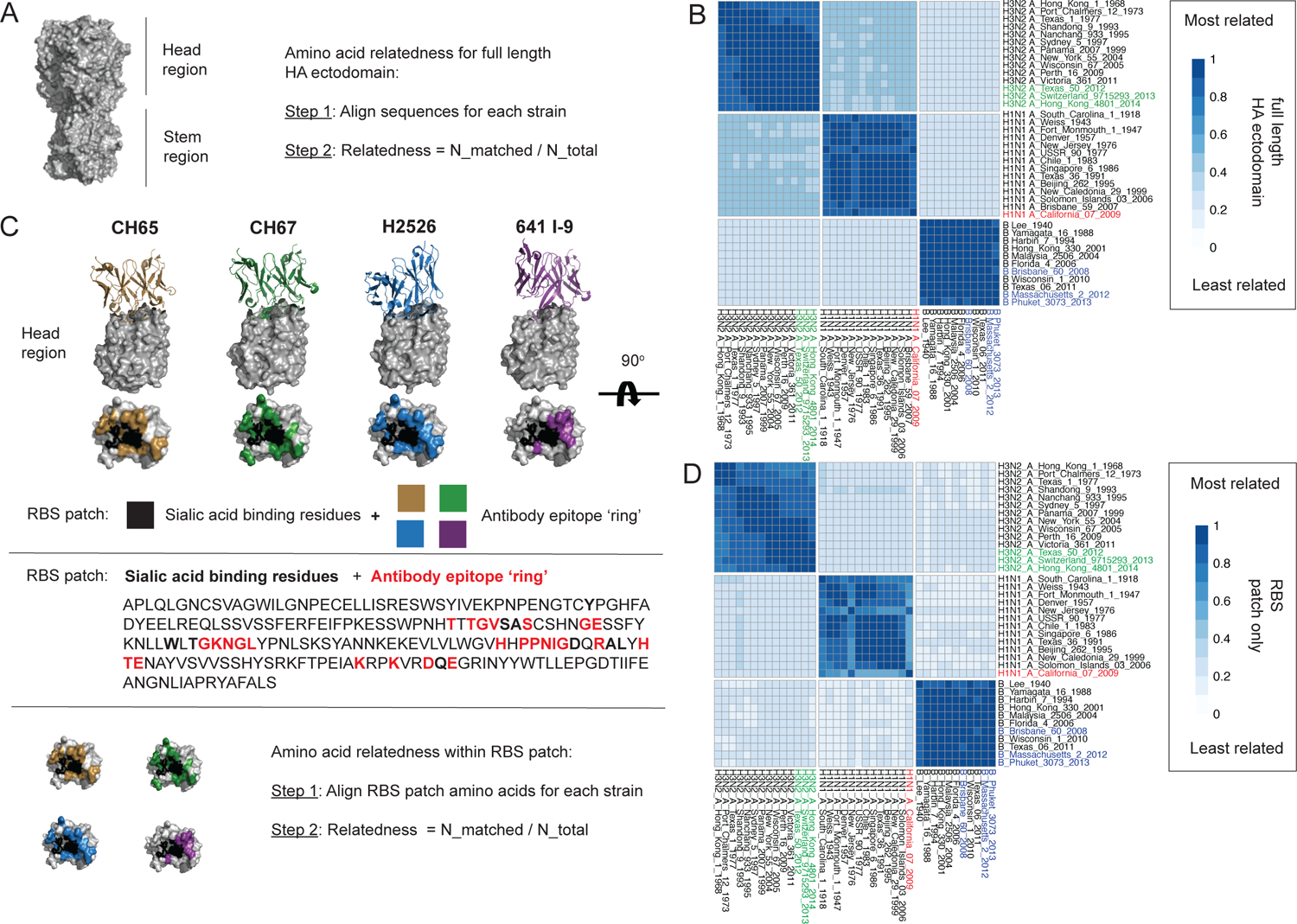
Divergent amino acid relatedness in the ectodomain and receptor binding site (RBS) patch of the pandemic influenza HA. (A) The HA ectodomain, where relatedness is calculated using the formula “N_matched / N_total” N_matched is the number of amino acids that match between the compared sequences and N_total is the total number of amino acids in the aligned sequence. (B) Heat map of HA ectodomain relatedness values for influenza A (H3N2, H1N1) and B viruses spanning >100 years (38 HA ectodomain sequences analyzed). (C) The RBS patch was structurally identified by four human bnAbs whose paratopes engage the RBS by mimicking cell surface sialic acid (CH67, CH67, H2526, 641 I-9) (Schmidt *et al*., 2015). We defined the RBS patch as the viral sialic acid binding residues (black) + the surrounding antibody epitope ‘ring’, collectively identified by the peripheral contacts made by the four bnAbs. Amino relatedness within the RBS patch is then calculated using the same formula except that the residues are now restricted to patch. (D) Heat map of HA RBS patch relatedness values for influenza A (H3N2, H1N1) and B viruses spanning >100 years (RBS patch sequences from the same 38 HA sequences as in B)

### Sequential vaccination with homologous pHA broadly boosts HAI

To define how sequential immunization with homologous HA impacts antibody scanning breadth in humans, we evaluated the HAI coverage across the diverse viral strains from our relatedness analysis (Figure 1B,D), as elicited by repeated (4x) inoculation with influenza vaccine containing the same H1N1 component (pHA) over a four year period (2013-2016) (Nunez *et al*., 2017) (Figure 2A). An important distinction from previous analyses of this dataset is that we now focus on the same individuals that were followed longitudinally across the four year period (n=27 individuals) (Data S1). In each year, a sample was obtained before and then twenty days after vaccination, and we first evaluated the fold change in HAI elicited against the virus panel (IBV, IAV H3N2, IAV H1N1) by each vaccine component within each year (Figure 2B,C). For H1N1, we find that pHA boosts HAI for the highly unrelated H1N1 strains, comparable to the HAI elicited for vaccine-matched A/California/07/2009; and also qualitatively similar to the HAI triggered by the H3N2 and IAB vaccine components (which are not novel pandemic hemagglutinins). These features were observed for full ectodomain or RBS patch (Figure 2B,C). Hence, immunization with homologous H1N1 pHA triggers elicitation of receptor-blocking antibodies against highly unmatched H1N1 viruses.

**Figure 2.**
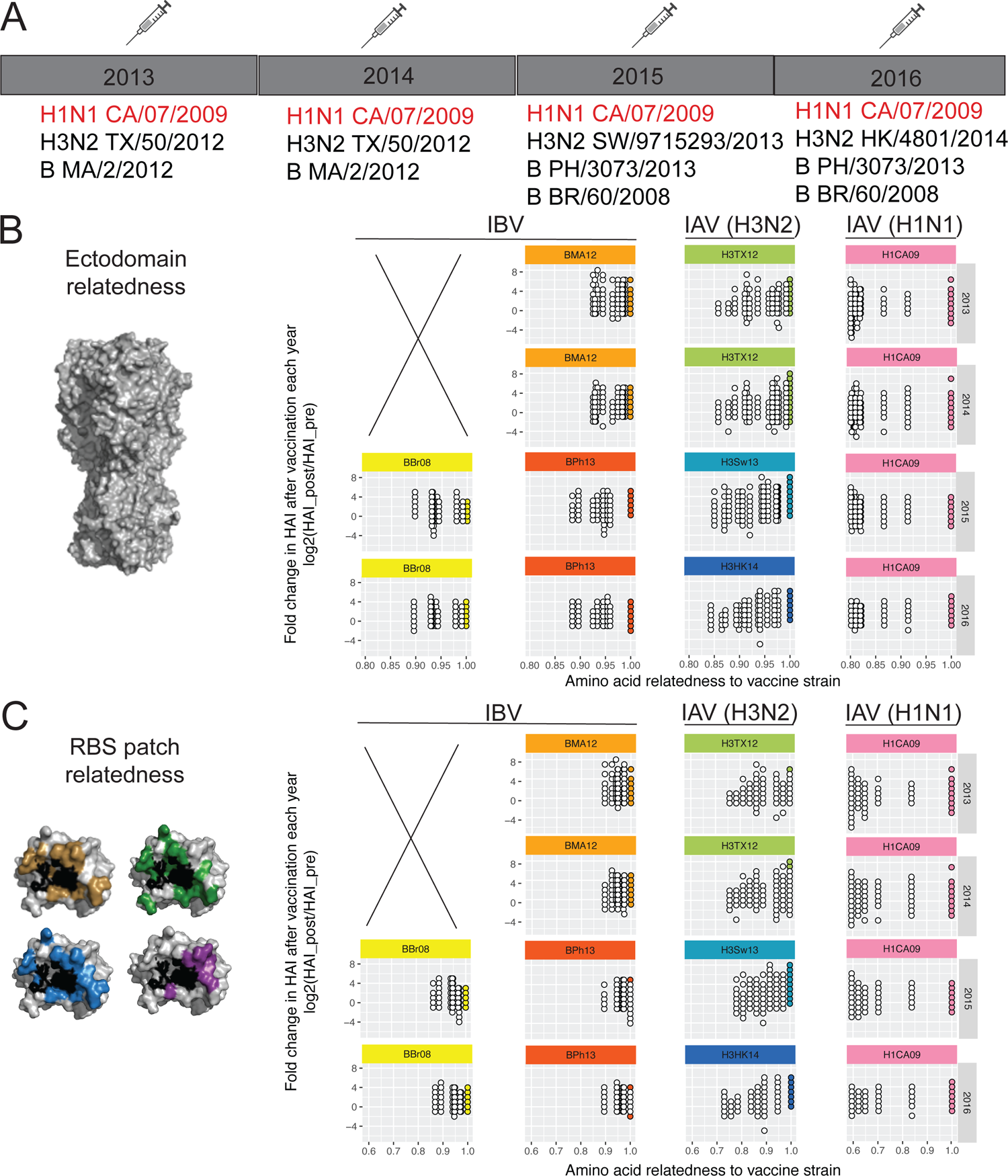
Sequential immunization with homologous pHA boosts highly unrelated H1N1 strains. (A) Four year influenza vaccine trial (Nunez, et al., 2017). We analyzed HAI elicited from 27 subjects that were longitudinally followed and immunized each year with the vaccine strains indicated. Notably these individuals received the same H1N1 component (pandemic A/California/07/2009) in each of the four years. (B) Fold change in HAI titer (pre vs 20 days post-vaccination) elicited each year and graphed as a function of HA ectodomain relatedness between the vaccine strain and the viruses within the HAI panels. Each dot is a single subject at the relatedness value: white dots are fold changes for strains from the virus panel; the colored dots indicate the vaccine-matched viral strain (relatedness = 1.00). (C) Same data as in (B) only now graphed as a function of RBS patch relatedness between the vaccine strain and the viruses within the panels.

### Sequential vaccination with homologous pHA broadens HAI with gradual kinetics in subjects that do not initially back-boost to historical strains

We next defined the kinetics of relatedness-independent broadening of H1N1 HAI over the four year period by graphing the fraction of responders vs non-responders (detectable vs non detectable boosting of HAI to each H1N1 strain) at each year (Figure 3A-C). Although pandemic HA will not be historically imprinted, memory recall of pre-existing immunity or ‘back boosting’ to historical strains would occur in response to the first antigen exposure (Akkaya et al., 2020; Henry *et al*., 2018; Nunez *et al*., 2017; Palm and Henry, 2019; Reusch and Angeletti, 2023; Turner et al., 2020) and cannot be ruled out in the first vaccine year (2013). For this reason we focused on the initial non-responders, who boost against pHA (and the other seasonal vaccine components, see Figure 2B,C) but do not simultaneously broaden/back-boost against historical H1N1 strains after immunization in Year 1, and by definition lack B cell memory that is recalled by pHA (Figure 3A-C). The subsequent reduction of these non-responders upon sequential vaccination with pHA in later years, identifies a separate vaccine broadening effect with slower kinetics (Figure 3A-C). In this effect, the proportion of non-responders to divergent H1N1 gradually decreases during the vaccine regimen, culminating in the near absence of non-responders in Year 4. This effect is seen when the subjects are not age stratified (Figure 3A) and when the subjects are divided into older and younger ages (>50 vs <38 years) (Figure 3B,C). The corresponding increases in the proportion of responders are also observed in these groups over the vaccine regimen (Figure S1). Collectively, these data indicate that within individuals that lack initial back-boosting, sequential exposure to pHA can broaden the RBS-directed antibodies against highly unrelated H1N1.

**Figure 3.**
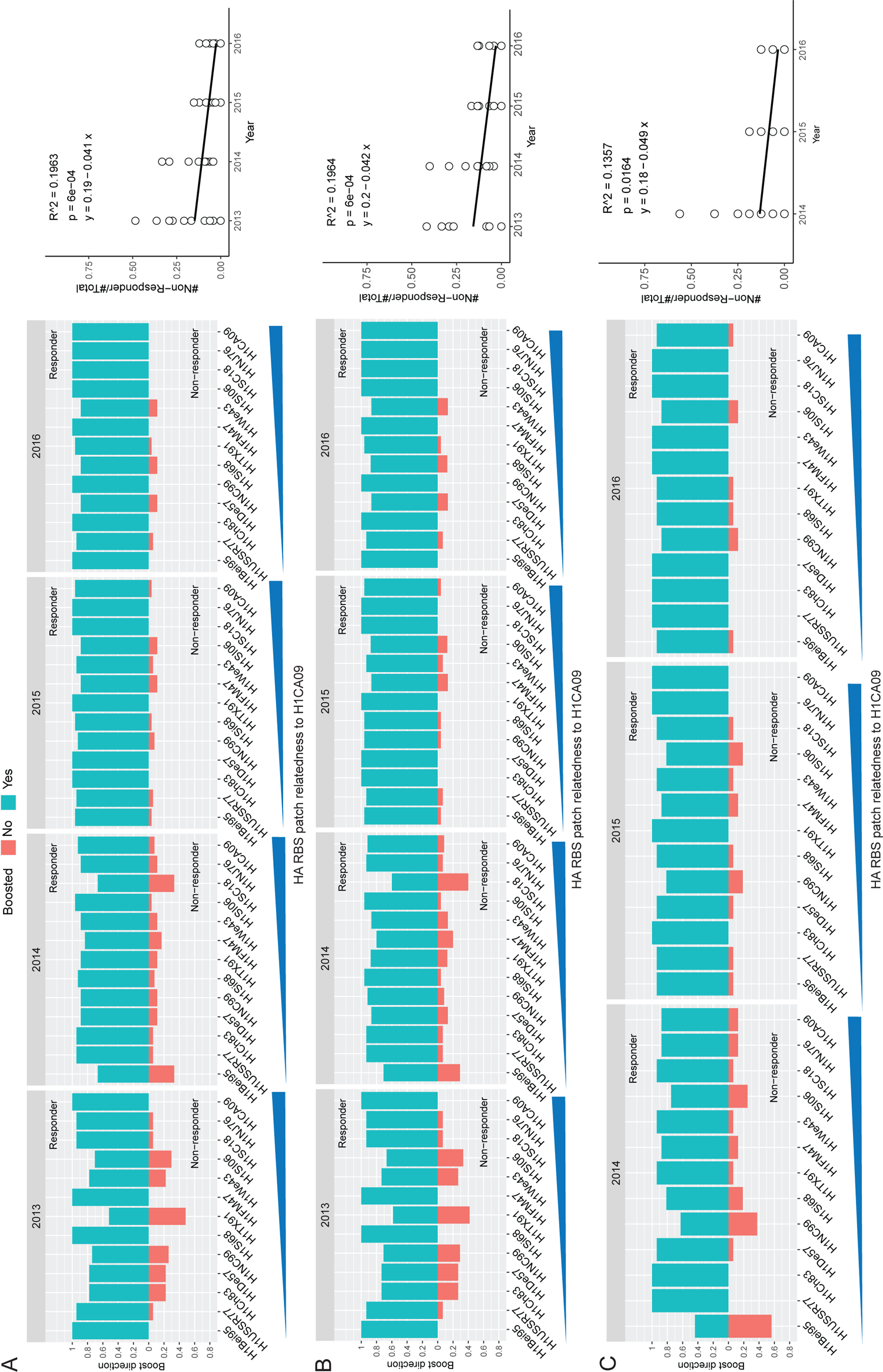
Sequential immunization with homologous pHA gradually broadens the response within individuals with no initial immune memory/recall to historical strains. Responders (green) versus non-responders (red) within each year is graphed for each H1N1 strain in the HAI panel. Responders are defined by having non-increasing fold changes of HAI titers (post-vaccination HAI titer / pre-vaccination HAI titer; i.e. fold change >1). Non-responders are defined by having decreasing fold changes of HAI titers (post-vaccination HAI titer / pre-vaccination HAI titer; fold change < 1). Because non-responders (red) do not back-boost against historical strains in the panel they, by definition, lack imprinted immunity to these viruses that is recalled by pHA. In the regression analyses each white dot denotes the proportion of non-responders for each viral strain. (A) Yearly response for all longitudinally analyzed individuals; at right is a linear regression of the proportion of non-responders against over the four year vaccine data (*P*=6e-04). (B) Data for subjects >50 years in age (*P*=6e-04, linear regression). (C) Data for subjects <38 years in age (*P=*0.0164, linear regression). See also Figure S1 for linear regression of the proportion of responders in each age group.

### A computational model to study the mechanistic origin of increased coverage following sequential immunization with homologous HA

To obtain mechanistic insights underlying the observed broadening of the antibody response in the absence of prior immune imprinting and back-boosting, we developed a computational model of the humoral immune response to RBS epitopes upon repeated vaccination. The model is principally an extension of our past work modeling humoral immune responses upon repeated vaccination with SARS-CoV-2 vaccine immunogens, but also builds on our other past studies (Amitai *et al*., 2020; Wang et al., 2015; Yang *et al*., 2023). The purpose of this model is not quantitatively fit clinical data, but to identify mechanistic principles that support the observations. Below, we outline the structure of the in silico model; mathematical and computational details are provided in Star Methods.

We first coarse-grained the HA RBS into three antibody epitopes (epitopes 1-3) on pHA (strain 1) and on two historical H1N1 strains (strains 2 and 3) (Figure 4A). In this model, a fraction *p_i_* of the germline B cells target epitope *i* and the immunodominance hierarchy is taken to be epitope 1 > epitope 2 > epitope 3. The immunodominant epitope on pHA (epitope 1) is heavily mutated as compared to strains 2 and 3. This is because the pandemic strain would escape previously immunodominant responses that target historical strains. We further assume that epitope 2 is relatively conserved between strains 1 and 2, but not conserved between strains 1 and 3; epitope 3 is relatively conserved between strains 1 and 3, but not strains 1 and 2 (Figure 4A). In this way, we model three historical strains that are different from each other, but share some similarities as would be expected for all H1N1 viruses.

**Figure 4:**
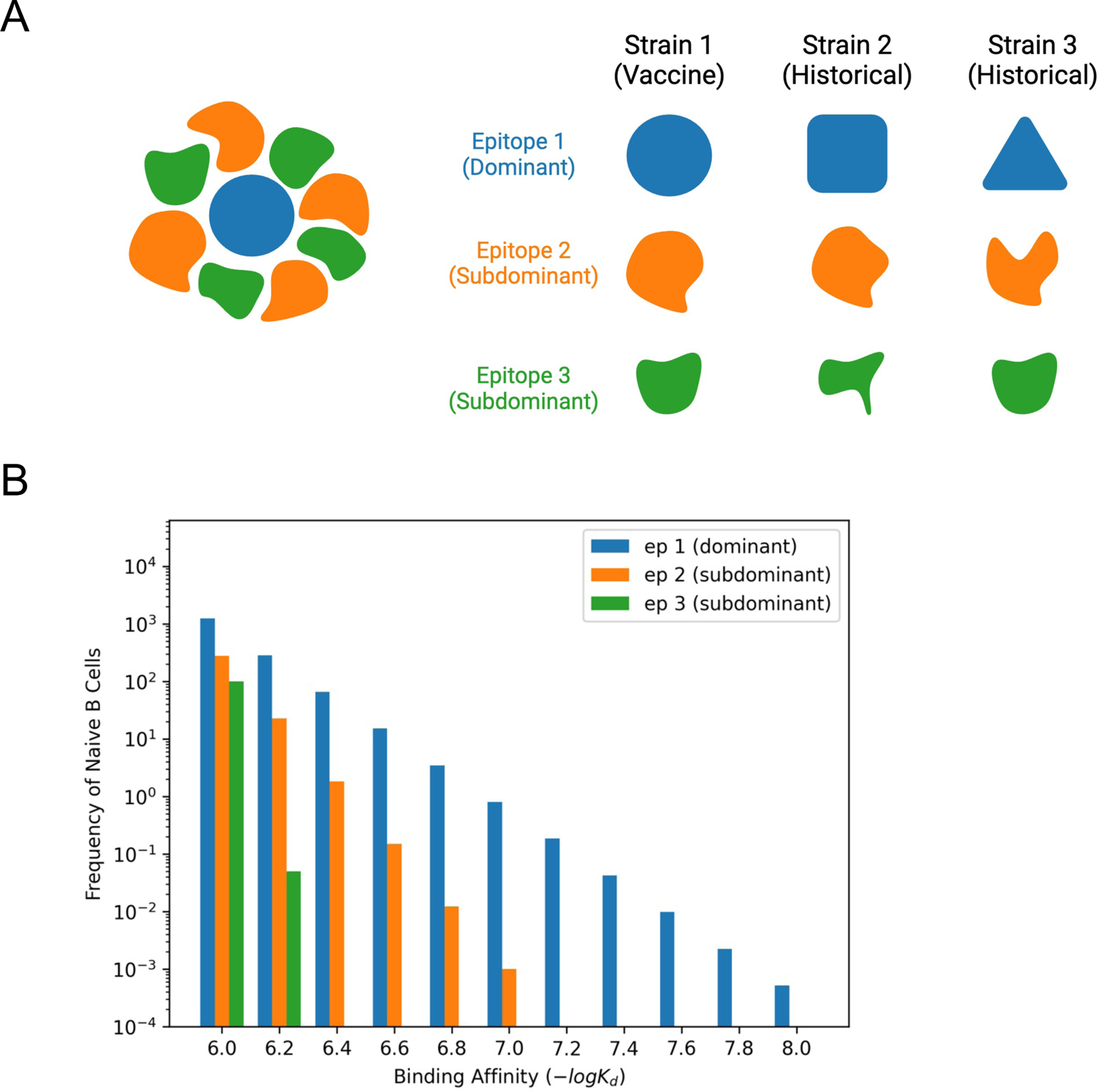
The influenza HA head is coarse-grained into three epitopes, that are perceived with different germline-endowed B cell affinities. (A) Diagram of epitope differences. In the right panel, the level of conservation of the three epitopes is depicted using different shapes (not very conserved) or similar shapes (relatively conserved) Epitope 1 (dominant epitope on pHA) varies is not conserved between the three variants. Epitope 2 (subdominant epitope) is relatively conserved between strain 1 (vaccine strain) and strain 2, but not between strains 1 and 3. Epitope 3 (another subdominant epitope) is conserved between strains 1 and 3, but not between strains 2 and 3. (B) Germline-endowed affinity distribution of naive B cells. More dominant epitopes have dominant than epitope 3. Here, the fractions of naive B cells pi targeting epitope *i* are p_1_ = a longer high-affinity tail. Epitope 1 is more dominant than epitope 2 and epitope 2 is more 0.8, p_2_ = 0.15, p_3_ = 0.05.

The immunodominance hierarchy of the three epitopes is reflected in the distribution of germline B cell affinities for antigen, an attribute that is important for B cell recruitment into GCs (Abbott and Crotty, 2020; Abbott et al., 2018; Amitai *et al*., 2020; Dosenovic et al., 2018; Sangesland and Lingwood, 2021). Detectable *g*ermline BCR affinities for antigen can range from 10^-7^ to 10^-4^ M (Feldman et al., 2021; Ronsard L, 2023; Sangesland, 2019; Sangesland et al., 2022), and a dissociation constant of ∼10^-6^ M is the estimate we use for the threshold for entry into GCs (Batista and Neuberger, 1998). Since high germline affinities are rare (Feldman *et al*., 2021; Kuraoka et al., 2016; Ronsard L, 2023; Sangesland, 2019; Sangesland *et al*., 2022), we consider the distribution of affinities to decay exponentially. More immunodominant epitopes constitute a larger fraction *p_i_* of germline B cells and exhibit a longer high-affinity tail (Figure 4B). We varied parameters that reflect the conservation of these epitopes and their relative immunodominance.

To then study humoral immune reactions to the different epitopes *in silico*, we modelled key steps that determine the antibody response pathway to protein antigens including: [**1**] antigen deposition on the surface of follicular dendritic cells (FDCs) (De Silva and Klein, 2015; Victora and Nussenzweig, 2022); [**2**] activation and entry of of naive B cells into germinal centers (GCs), affinity and T helper-cell-driven selection within GCs (De Silva and Klein, 2015; Victora and Nussenzweig, 2022; Young and Brink, 2021), and differentiation into memory B cells and plasma cells (Akkaya *et al*., 2020; Crotty, 2015; Palm and Henry, 2019); [**3**] relatively rapid expansion and differentiation of memory B cells into short-lived plasma cells during the recall responses which occur outside GCs or in extra germinal center locations (EGCs) (Moran et al., 2018; Van Beek et al., 2022).

A set of differential equations was used to model the dynamics of antigen deposition and presentation on FDCs (see Star Methods for details). Circulating antibodies can bind to soluble antigen to form immune complexes (ICs). For the first immunization, we assume that only weakly binding circulating IgM antibodies are available for binding to the antigen and forming ICs, which are then deposited on FDCs. ICs deposited on FDCs are longer-lived than soluble antigen. In the first few days after vaccination, soluble antigen rapidly decays (Aung et al., 2023; Martin et al., 2021). Therefore, the weakly binding IgM antibodies can deposit relatively small amounts of ICs on FDCs before soluble antigen decays. For subsequent immunizations, stronger binding antibodies elicited by the previous immunization are available to bind antigen and form ICs that are deposited on FDCs before soluble antigen decays. The differential equations that describe these processes are coupled to an agent-based simulation of the stochastic processes that occur in GCs and EGCs to produce memory B cells and antibodies. The stochastic simulations also model GC entry of B cells.

In the stochastic agent-based simulations of GCs and EGCs, each B cell is an agent and the probabilities of its activation, selection, proliferation, mutation, and differentiation are calculated at each time step (0.01 days). Our model accounts for the following immunological principles and factors in the activation and selection of B cells: GC B cells internalize antigen based on their binding affinities to epitopes in the vaccine strain (Batista and Neuberger, 1998; Fleire et al., 2006); the amount of antigen internalized grows with the antigen binding free energy (or affinity) and saturates above a threshold affinity (Foote and Eisen, 1995; 2000); and individual B cells compete for subsequent T cell help to promote B cell survival. Among the B cells that are positively selected in the GC, some stochastically exit the GC and differentiate into either plasma or memory B cells (Akkaya *et al*., 2020; Crotty, 2015; Palm and Henry, 2019). The majority of positively selected B cells are recycled for mutation-selection cycles and they proliferate and undergo somatic hypermutation (SHM) (Collins and Jackson, 2018; De Silva and Klein, 2015; Glanville et al., 2009; Li et al., 2004; Mesin et al., 2016; Victora and Nussenzweig, 2012; 2022). SHM is responsible for affinity-changing mutations, though it also leads to apoptosis or no affinity change with different probabilities (Amitai *et al*., 2020; Wang *et al*., 2015; Yang *et al*., 2023; Zhang and Shakhnovich, 2010). Based on data from experiments on affinity changes upon mutations at protein-protein interfaces, the change in affinity due to mutation is drawn from a log normal distribution with only 5 % of mutations being beneficial (Kumar and Gromiha, 2006; Zhang and Shakhnovich, 2010). A summary of the mathematical details of the steps described above is provided in Star Methods.

In our model, memory cells are stochastically selected in an affinity-dependent way and expanded in EGCs via the same processes as in GCs, except that there are few to no mutations (none in our model) (Moran *et al*., 2018; Van Beek *et al*., 2022; Yang *et al*., 2023). B cells exiting EGCs differentiate into antibody-secreting plasma cells with a probability of 0.6, because experimental data shows that 60 % of new proliferating memory B cells differentiate into plasma cells (Moran *et al*., 2018). These plasma cells produce antibodies at rates estimated from experiments (Goel et al., 2021; Muecksch *et al*., 2022). Our model also incorporates epitope masking in which circulating antibodies specific for a given epitope can enter ongoing GCs and EGCs and compete with B cells specific for the same epitope (Bergstrom et al., 2017; Schaefer-Babajew et al., 2023; Tas et al., 2022; Zarnitsyna et al., 2016).

While GC and EGC processes are driven by the vaccine antigen (strain 1), we also track the affinities of the resulting memory B cells and antibodies for strains 2 and 3 as well. A B cell’s affinity for each strain depends on its initial affinity and the affinity-changing mutations that occur within the GC. The size of affinity-changes upon mutation for the three epitopes on described by a parameter, ρ, that determines a covariance matrix. This parameter is related to the different strains are drawn from correlated log-normal distributions. The level of correlation is level of conservation and amino acid relatedness between the strains for the B cell’s target epitope. For instance, if 70% of the amino acids in an epitope are shared between strains 1 and 2, strain 1 are beneficial for strain 2 as well. The value of ρ, for B cells targeting epitope 2 is high we can approximate that ∼70% of the beneficial mutations for B cells targeting this epitope in between strains 1 and B, but low between strains 1 and 3. The value of ρ, for B cells targeting epitope 3 is high between strains 1 and 3, but low between strains 1 and 2. The value of ρ, for B cells targeting epitope 1 is low between strain 1 and strains 2 and 3 as it is poorly conserved in historical strains. We vary the values of the parameter, ρ,, and study the effects.

For every immunization, we simulate 200 GCs and 1 EGC. Ten different simulations are carried out for any given condition and the results shown are averages over these simulations. The parameters used in the simulations are provided in Table S2.

### Booster shots of homologous pHA provide increasing coverage of historical strains through pathways that characterize the humoral response

Figure 5A shows the results of our simulations for antibody titers elicited against strain 1 (pHA) and the two historical strains (strains 2 and 3) after each of four immunizations with strain 1. The titers are calculated based on the affinity and number of antibodies that target the epitopes in each strain. After the first immunization, significant titers of antibodies are generated only against the dominant epitope of strain 1. However, after the second immunization the titers are boosted against all strains, including strains 2 and 3. Continued boosting with pHA continues to amplify heterologous coverage, even when the homologous boosting titer has plateaued.

**Figure 5.**
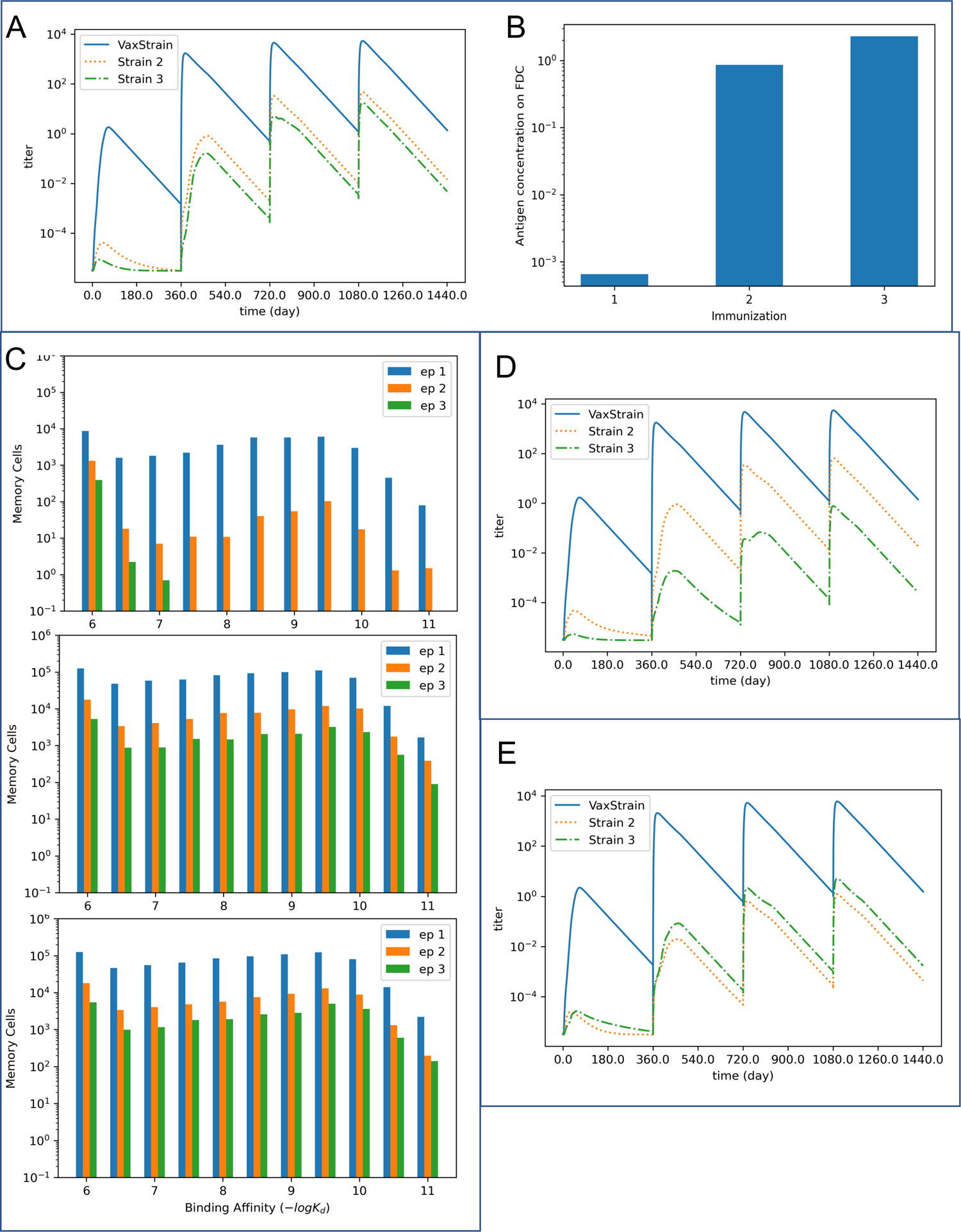
Antibody broadening via feedback loops within the humoral response. (A) The antibody titers against both the vaccine strain and historical strains (strains 2 and 3) increase over four immunizations. The antibodies are produced by plasma cells from both the GCs and the EGCs. Antibody coverage increases first for strain 2 (after the second immunization) and strain 3 is engaged after the third immunization. In this simulation, the initial fractions of B cells *p*_i_ that target epitope *p*_i_ are *p*_1_ = 0.8, *p*_2_ = 0.15, *p*_3_ = 0.05. The conservation *p*_12_ of epitope 2 between strains 1 and 2 and the conservation *p*_13_ of epitope 3 between strains 1 and 3 are both equal to 0.95. (B) The expansion of pathogen-specific IgG antibodies from the first immunization significantly increases the antigen concentration on the FDC in the second immunization. This allows lower-affinity B cells that target subdominant epitopes to enter GCs and undergo affinity maturation. The antigen concentration on the FDC slightly increases from the second to the third immunization, allowing more B cells that target the subdominant epitopes to enter GCs and undergo affinity maturation. (C) The distribution of memory cells produced in the GCs during the first three immunizations. Upon subsequent antigen exposure, these memory cells are selected and expanded in EGCs. Thus, they contribute significantly to circulating antibodies and increased titers during subsequent immunizations. The first immunization primarily produces memory cells that target the dominant epitope (epitope 1), along with some memory cells targeting epitope 2. The second and third vaccinations produce an overall greater number of memory cells overall bearing higher affinity for the subdominant epitopes (epitopes 2 and 3) than the first immunization. (D) Strain 3 is engaged less potently when the initial fractions of B cells *p*_i_ that target epitope *p*_i_ are *p*_1_ = 0.8, *p*_2_ = 0.18, *p*_3_ = 0.02. (E) The titers against strain 2 are conservation *p*_12_ of epitope 2 between strains 1 and 2 is 0.7 while the conservation *p*_13_ of lower than titers against strain 3 when the conservation of epitope 2 is decreased. Here the epitope 3 between strains 1 and 3 is kept at 0.95. Other values of *p*_12_ are explored in Figure S2. The fractions of B cells *p*_i_ that target epitope i are the same as those in Figure 5A.

Experiments show that injected soluble antigen decays relatively rapidly (Aung *et al*., 2023; Martin *et al*., 2021; Tam et al., 2016). The vaccine antigen is the pandemic strain and the individuals studied do not initially backboost responses to historical strains. Therefore, upon the first immunization only generic circulating IgM antibodies, with low affinity for the antigen, are available to form the immune complexes needed for antigen deposition on FDCs (see computational methods section). Thus, our computational results show that very little antigen is deposited on FDCs after the first immunization with a new antigen (Figure 5B).

The germline B cells targeting the immunodominant epitope on strain 1 are more abundant and generally have higher affinities than the B cells engaging the subdominant epitopes. When antigen available on FDCs is low after the first immunization, the greater abundance and affinities of these germline B cells confers an especially strong advantage to them in entering GCs compared to B cells targeting subdominant epitopes. Furthermore, they also are much more likely to dominate GC reactions during affinity maturation. Thus, the high affinity memory B cells generated after the first immunization predominantly target the immunodominant epitope (Figure 5C, top panel).

After the second immunization, the EGCs facilitate the production of antibodies that engage strain 1, but not the historical strains. This is because the available memory B cells after the first immunization largely target the immunodominant epitope that is not conserved in the historical strains (Figure 5C, top panel). Thus, for many days after the second immunization, significant titers of antibodies that can target historical strains are not elicited (Figure 5A). However, secondary GCs also form during this time. Upon the second immunization, higher affinity IgG antibodies specific for the antigen that were generated during the first immunization are available to bind to the antigen and deposit it on FDCs before the antigen is degraded (Figure 5B). Higher amounts of deposited antigen allow lower-affinity germline B cells that target sub-dominant epitopes to enter the GC and be positively selected. As affinity maturation proceeds, these GC B cells acquire higher affinity to the subdominant epitopes that are relatively conserved between strain 1 and the historical strains, and generate high affinity memory B cells (Figure 5C middle panel). The plasma cells produced by the GCs produce antibodies that engage strains 2 and 3 (particularly strain 2 since epitope 2, the second most dominant epitope, is relatively conserved between strains 1 and 2).

After the third immunization, memory cells generated after the second immunization that target epitopes 2 and 3 with high affinity are also selected based on their affinities and expanded in the EGC. This results in further amplification of antibodies that target epitopes 2 and 3 (Figure 5A). Antigen deposition on FDCs is also somewhat elevated after the third immunization (Figure 5B). So, strain 3 is engaged more potently and this increases the relative number of memory B cells targeting epitope 3 compared to epitope 2 among the memory B cells generated in GCs after the third immunization (Figure 5C, bottom panel). As a consequence, the difference between the titers produced against epitopes 2 and 3 is further decreased after the fourth immunization (Figure 5A). However, the overall improvements to antibody titers are minor because antigen presentation in the GCs and expansion of memory B cells in EGCs is similar to that after the third immunization.

The relative coverage of strains 2 and 3 upon repeated vaccination depends upon the relative immunodominance of epitopes 2 and 3 in the pool of germline B cells. To test the effects of modifying the immunodominance hierarchy, we increased the fraction of germline B cells that target epitope 2, and decreased the fraction that target epitope 3. This enhances the immunodominance of epitope 2 over epitope 3. Consequently, as shown in Figure 5D, the responses to epitope 3 and to historical strain 3 in which epitope 3 is conserved are less potent compared to the results shown in Figure 5A.

The relative coverage of historical strains also depends on the conservation of subdominant epitopes between the historical and immunizing strains. In Figure 5A, the conservation of epitope 2 between strains 1 and 2 and of epitope 3 between strains 1 and 3 are the same. The results show that higher titers are elicited against strain 2 than strain 3, which aligns with the immunodominance of epitope 2 over epitope 3. However, it is possible that a more immunodominant epitope is less conserved as viruses mutate to avoid immune detection (Altman *et al*., 2018; Angeletti et al., 2017; Angeletti and Yewdell, 2018). Thus, we examined the effects of reducing the conservation of epitope 2 while fixing the conservation of epitope 3. If the conservation of epitope 2 between strains 1 and 2 is decreased below a critical value (see Figure S2), the advantage of epitope 2 due to a more favorable germline distribution is outweighed by weaker conservation. In Figure 5E, epitope 2 is more weakly conserved than in Figure 5A (but the germline immunodominance hierarchy is kept the same), resulting in lower titers against strain 2 than against strain 3.

### Epitope masking enhances the generation of antibodies that can engage historical variants upon boosting with unmatched homologous HA

Circulating antibodies can enter GCs and bind to their corresponding epitopes on antigen presented on FDCs (Schaefer-Babajew *et al*., 2023; Tas *et al*., 2022). This masking of an epitope by soluble antibodies lowers the effective amount of antigen available to GC B cells targeting the same epitope, reducing their competitive fitness within the GC. This property of epitope masking by circulating antibodies can be applied to regulate the GC participation of naive B cells according to epitope specificity and the competitive environment for GC B cells (Schaefer-Babajew *et al*., 2023; Tas *et al*., 2022; Yang *et al*., 2023). Our computational model shows that after the second immunization, most of the circulating antibodies bind to the dominant epitope. This is because most of the memory B cells produced after the first immunization are directed against the dominant epitope (Figure 5C and 6C), and these memory cells are rapidly expanded in EGCs after the second immunization to generate the corresponding antibodies. Accordingly, we find that the entry and selection of subdominant-targeting B cells during affinity maturation are enhanced by including the effects of epitope masking which generates more memory cells that target the subdominant epitopes (Figure 6C and 6D). This results in larger antibody titers targeting strains 2 and 3 if epitope masking is included (Figure 6A). Likewise, after the third immunization, the antibodies from the second vaccination begin to mask epitope 2, promoting the generation of memory B cells and antibodies targeting epitope 3 (Figure 6). This feature of increasing antibody titers against the historical strains is observed when we consider two cases of epitope masking (Figure 6A and 6B): [1] where the three epitopes under consideration are non-overlaping; and [2] where the epitopes overlap with each other. The first case shows the largest effect of epitope masking. In the second case, epitope masking effects remain significant, even if we consider a modest level of epitope overlap (e.g. 30% between epitope 1 (dominant) and epitope 2 (subdominant); and 30% between epitope 1 (dominant) and epitope 3 (subdominant)).

**Figure 6.**
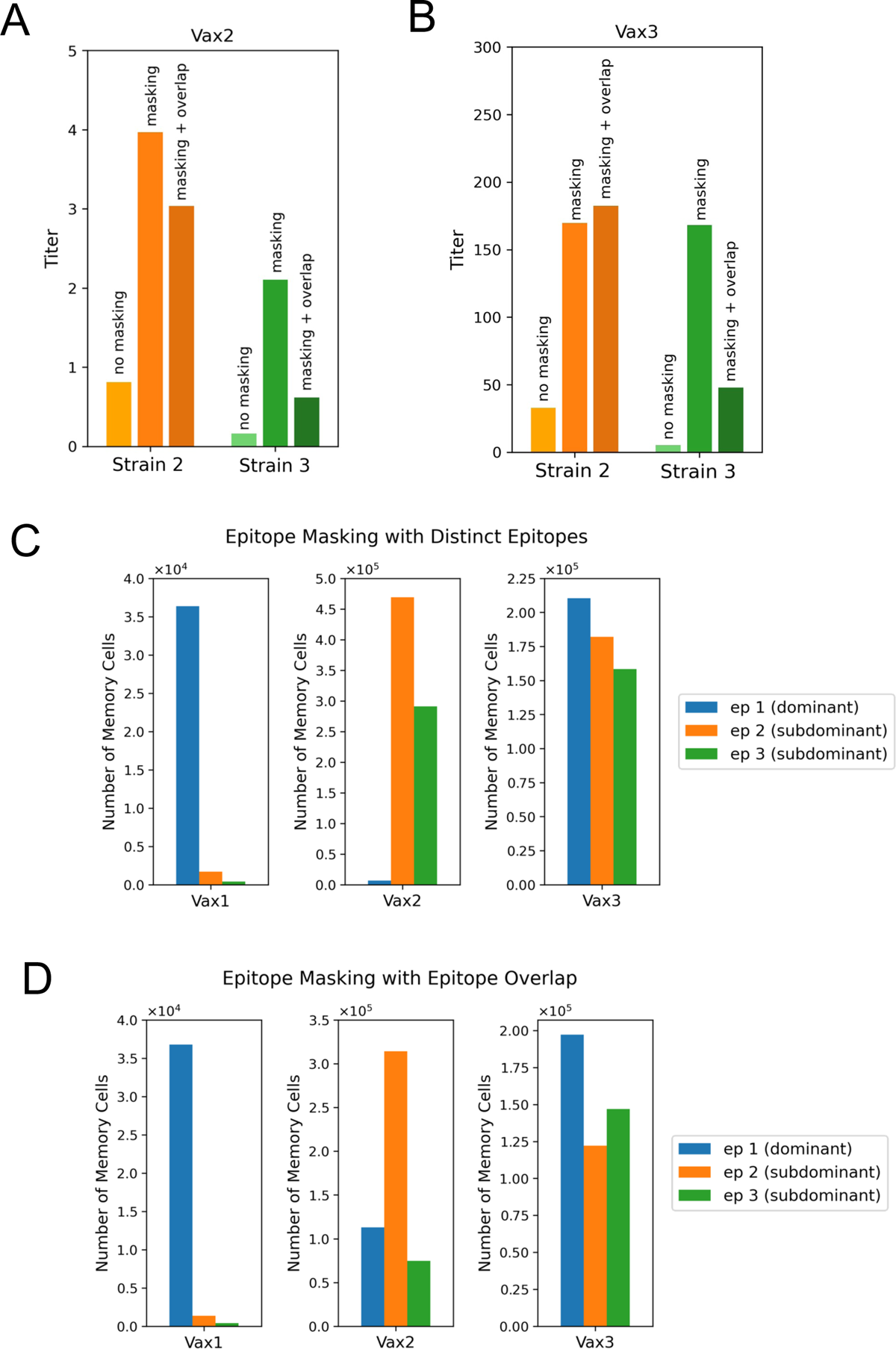
Regulation of antibody broadening through epitope masking. (A) Maximum antibody titers for the historical strains after the second vaccination, with and without epitope masking. Two cases are considered when there is epitope masking: [1] the epitopes are absolutely distinct; [2] the epitopes can overlap with each other. The second case is shown here, wherein there is 30 % overlap between epitope 1 (dominant) and epitope 2 (subdominant) and between epitope 1 (dominant) and epitope 3 (subdominant). Masking increases the titers against historical strains, even when there is some overlap between the dominant and subdominant epitopes. (B) Maximum antibody titers for the historical strains after the third vaccination, with and without masking. After the third vaccination, titers for Variant 2 with epitope masking are higher when there is epitope overlap than when the epitopes are distinct. (C) Relative number of memory cells produced with epitope masking. Epitopes are considered to be fully distinct. The epitope that is most targeted by the memory cells is also masked the most after the subsequent immunization. The dominant epitope is targeted most by Vax 1 memory cells and is masked the most in the second immunization. The orange subdominant epitope (epitope 2) and green subdominant epitope (epitope 3) are both relatively well targeted by Vax 2 memory cells. However, the subdominant epitopes are also masked during the third immunization, so the subdominant epitopes lose their advantage compared to the dominant epitope in the affinity maturation process after Vax 3. (D) Relative number of memory cells produced with epitope masking and overlap. The epitope that is most targeted by the memory cells is masked the most in the subsequent immunization. The dominant epitope is targeted most by Vax 1 memory cells and is masked the most in the second immunization. The orange subdominant epitope (epitope 2) is targeted most by Vax 2 memory cells, although more memory cells target the dominant epitope than when the epitopes are fully distinct. Due to the masking of epitope 2 in the third immunization, the dominant and green subdominant epitope (epitope 3) are both relatively well targeted by Vax 3 memory cells.

Epitope overlap between dominant and subdominant epitopes generally decreases titers against historical strains compared to fully distinct epitopes. However, after the third vaccination the antibody response titer against strain 2 is higher when there is epitope overlap (Figure 6B, right panel). This is because the second immunization produces more memory cells targeting the dominant epitope when there is epitope overlap (Figure 6D). These memory cells are rapidly expanded in EGCs and so antibodies mask the dominant epitope more strongly during the third immunization when the epitopes overlap. This leads to the production of more high-affinity B cells that target epitope 2 after the third immunization, and subsequently higher titers against strain 2.

## Discussion

Our results reveal broadening of antibody coverage following repeated immunization with homologous pHA. This occurred over a period when the subjects had limited imprinting to this vaccine strain, and the process occurred gradually within individuals that did not initially ‘back-boost’ against historical H1N1 strains. These findings are consistent with sequential exposure to SARS-CoV-2 mRNA vaccines, where repeated immunization with the homologous Wuhan vaccine strain eventually elicits non-imprinted coverage against highly unmatched Omicron-lineages of SARS-CoV-2 (Garcia-Beltran *et al*., 2022; Muecksch *et al*., 2022; Schmidt *et al*., 2022). Our computational results describe how aspects of the humoral response (enhanced antigen presentation on FDCs and epitope masking) are a likely origin of eliciting non-imprinted antibody breadth to influenza HA. These mechanisms are consistent with the immune reactions that broaden antibodies after boosting with the homologous SARS-CoV-2 vaccine (Yang *et al*., 2023). Collectively, our results point to a general feature of the humoral response that intrinsically broadens antibodies against unmatched/diversified antigen targets upon repeated vaccination with the same novel antigen.

This principle of preserving diversified or ‘non-homogenized’ antibody output during on-going immune reactions has emerged as an important theme for germinal center and memory B cell responses to protein antigens (de Carvalho et al., 2023; Hagglof et al., 2023; Kuraoka *et al*., 2016; Mesin *et al*., 2016; Mesin et al., 2020; Radmacher et al., 1998; Ronsard L, 2023; Sabouri et al., 2014; Silver et al., 2018; Tas et al., 2016; Van Beek *et al*., 2022). Our *in silico* results would suggest that intrinsic broadening of the antibodies generated in response to homologous antigen is an extension of this fundamental principle, enabling polyspecific responses and broad B cell reactivities that are tuned by antigen presentation and epitope masking effects.

Operationally, it will be important to integrate this information to define the number of homologous immunizations needed to elicit a given level of vaccine coverage of historical or new strains. Our simulations indicate antibody broadening depends on the interplay of several key factors including: the precursor frequencies of germline B cells targeting these epitopes, the affinities of germline B cells for their target epitopes, and the conservation of shared epitopes within the RBS. Precursor frequencies and germline affinities determine the B cell pool’s initial affinities toward target strains—historical or emergent—while conservation of shared epitopes governs how those affinities change during affinity maturation in response to selection of GC B cells for their affinities to the immunizing strain. Simulation results show that the effects of the precursor frequencies and germline affinities can compete with the effects of epitope conservation in determining the coverage of a particular strain.

Our results also suggest that the germline B cell-mediated effects noted above are particularly important in earlier stages of the sequential vaccine regimen. But once higher affinity antibodies circulate (and recall of B cell memory becomes operational), epitope masking effects and enhanced antigen deposition on FDCs further modulate the immunodominance hierarchy to favor responses to sub-dominant epitopes. While our model does not account for memory B cells seeding secondary GCs, our previous work indicates that if more memory B cells enter secondary GCs the evolution of broadened antibody responses is diluted (Yang *et al*., 2023).

In all cases, our simulations recapitulate the clinical vaccine results in a qualitative manner. This owes to our development of a simplified model for the vaccine antigen, its relation to historical strains, and the characteristics of the germline B cells that engage RBS epitopes. While recapitulation of antibody broadening through simplified computational models points to fundamental/core features of adaptive immunity, more comprehensive epitope mapping of the B cell responses to the RBS targets (structure, engagement affinity, immunodominance patterns) will be needed to assemble more complete prediction maps to guide vaccine design.

In summary, we describe a phenomenon that defies the long-held view that sequential immunization with homologous influenza HA antigens serves to only/principally reinforce the boosting of antibodies with narrow coverage (Henry *et al*., 2018; Krammer *et al*., 2018; Krammer and Palese; Sangesland and Lingwood, 2021). Previous B cell epitope prediction algorithms have failed to computationally delineate and accurately predict patterns in the humoral response (Mahanty et al., 2015; Rockberg and Uhlen, 2009; Sela-Culang et al., 2013; Van Regenmortel, 2002; 2011), but by modeling the key immunological steps that underscore adaptive immune reactions to protein antigen (Akkaya *et al*., 2020; Crotty, 2015; De Silva and Klein, 2015; Palm and Henry, 2019; Victora and Nussenzweig, 2022; Young and Brink, 2021), we recapitulate the clinical findings and describe mechanisms that underlie them. This broadening effect has thus far only been observed in human antibody responses to vaccination and we suggest that it may further inform universal vaccine approaches (Caradonna and Schmidt, 2021; Krammer *et al*., 2018; Sangesland and Lingwood, 2021; Wei *et al*., 2020). Immunologically, we suggest that antibody broadening reflects an in-built feature of continued B cell diversification, a principle that will insure antibody complementarity to hypervariable antigen targets.

### Limits of the Study

This work lays the groundwork to experimentally define factors identified as significant for antibody broadening *in vivo*, including epitope conservation, epitope masking, and antigen presentation on FDCs. While the model predicts that epitope conservation impacts titers against historical strains, the data examined in this study do not show a clear trend between amino acid similarity with the vaccine strain and the titer levels or coverage kinetics among H1N1 strains. Exploring alternate experimental metrics for epitope conservation instead of amino acid similarity, along with examination of diverse sets of clinical data, can improve the model and lead to better congruence between model predictions and data. In addition, experimentally defining the relative contribution of epitope masking and antigen presentation on FDCs will necessate vaccine/immune challenge models where these parameters can be measured individually and that accurately reflect human humoral response features (Aung *et al*., 2023; Schaefer-Babajew *et al*., 2023; Tas *et al*., 2022; Zarnitsyna *et al*., 2016). In humans, an accurate description of individual imprinted B cell memory to influenza HA will also be needed to predict the broadening activity of homologous influenza vaccine regimen, in lieu of another pandemic influenza virus strain that does not back-boost to historical strains.

## Data Availability

All data produced in the present study are available upon request to the authors. The raw data is also provided in Data S1.

## Acknowledgements

MT and AKC acknowledge significant discussions with L. Yang. DL and YD thank the Lingwood lab for helpful feedback. The authors acknowledge support form NIH grants [U19AI057229 (AKC); AI155447, AI137057, and AI153098 (DL) AI146779, AI089618 (AGS)]; NIH contract 75N93019C00050 (AGS); NIH contract 75N93019C00052 (D.L.); as well as support the Ragon Institute of MGH, MIT & Harvard. YD was supported by a Schwartz AI initiative. TMR is also supported, in part, as a Georgia Research Alliance Eminent Scholar. The authors would also like to acknowledge Harvard CFAR for ongoing support through P30 AI060354.

## Author contributions

Conceptualization, DL, AKC; Investigation, YD, MT; Writing – Original Draft, YD, MT, AKC, DL.; Writing – Review and Editing, all authors; Funding Acquisition, DL, AKC; Supervision, DL, AKC.

## Declaration of interests

DL reports SAB membership for Metaphore Bio (a Flagship company), Tendel Therapies, and Lattice Therapeutics Inc. AKC is a consultant (titled “Academic Partner”) of Flagship Pioneering, a consultant and member of the Strategic Oversight Board of Apriori Bio (a Flagship company), and a consultant and SAB member of Metaphore Bio.

## STAR METHODS

### LEAD CONTACT AND MATERIALS AVAILABILITY

#### Lead Contact

Further information and requests for reagents should be directed to and will be fulfilled by Daniel Lingwood and Arup Chakraborty.

#### Materials Availability

There are no restrictions on the availability on the materials used in this study.

#### Data and Code Availability

All longitudinal HAI values used in this study are provided in Data S1. All original code and data files for the computational results have been deposited at https://github.com/mtang17/flu and are publicly available.

## EXPERIMENTAL MODEL AND SUBJECT DETAILS

We evaluated the HAI titers elicited by a clinical influenza vaccine that was sequentially immunized (4x) over a four year period (2013-2016) and contained the same H1 component (A/California/7/2009) in each of the vaccine years (Nunez *et al*., 2017). An important distinction from the previous analysis is that we focused only on individuals that were longitudinally sampled across the four year period (n=27 individuals; subjects are grouped into two age categories: above 50 years old and below 38 years old, see Data S1). In each year a blood sample was obtained before and then twenty days after vaccination. HAI titers for viruses spanning 100 years of influenza evolution were measured in each of these samples (Nunez *et al*., 2017) (see Data S1 and Table S1). The H1N1 viral panel for HAI comprised*: A/South Carolina/1/1918, A/Weiss/JY2/1943, A/Fort Monmouth/1/1947, A/Denver/1/1957, A/New Jersey/6/1976, A/USSR/90/1977, A/Chile/1/1983, A/Singapore/6/1986, A/Texas/36/1991, A/Beijing/262/1995, A/New Caledonia/20/1999, A/Solomon Island/3/2006, A/Brisbane/59/2007, A/California/07/2009*. The H3N2 vaccine panel for HAI comprised: *A/Hong/Kong/1/1968, A/Hong/Kong/4801/2014,A/Nanchang/933/1995, A/New/York/55/2004, A/Panama/2007/1999, A/Perth/16/2009, A/Port/Chalmers/12/1973, A/Shandong/9/1993, A/Switzerland/9715293/2013 A/Sydney/5/1997, A/Texas/1/1977, A/Texas/50/2012, A/Victoria/361/2011, A/Wisconsin/67/2005.* The IBV viral panel for HAI included: *B/Brisbane/60/2008, B/Florida/4/2006, B/Harbin/7/1994, B/Hong/Kong/330/2001, B/Lee/1940, B/Malaysia/2506/2004, B/Massachusetts/2/2012, B/Phuket/3073/2013, B/Texas/06/2011, B/Wisconsin/1/2010, B/Yamagata/16/1988*

## METHOD DETAILS

### Amino acid relatedness in HA ectodomains or within the RBS patch

Amino acid sequences of HA ectodomains from the different strains used in HAI were obtained from Genbank (https://www.ncbi.nlm.nih.gov/genbank/) or GISAID (https://gisaid.org/) (Table S1). The amino acid positions comprising the HA RBS patch was defined by the structures of four human broadly neutralizing RBS-directed antibodies, each in co-complex with HA (Schmidt *et al*., 2015). We used this information to define amino acid relatedness between the HA glycoproteins of all the influenza viruses used in our HAI panel. Pairwise relatedness was defined for both full length HA ectodomain and for the RBS patch. In both cases, amino acid sequence relatedness was obtained by first aligning two amino acid sequences and then computing the ratio of matched amino acid counts over the total amino acid counts in the aligned sequences. Heatmaps visualizing the pairwise amino acid sequence relatedness values were relatedness is represented as epitope conservation with the parameter ρ, in the computational graphed using “pheatmap” function from R package “pheatmap” (version 1.0.12). Amino acid model (see modeling section below).

### Analysis of HAI titers in relation to amino acid relatedness

We constructed dot plots to visualize the relationship between the fold change of HAI titers to the individual viruses in relation to their amino acid relatedness (for both the HA ectodomain and RBS patch) to the vaccine strain used in each year. In the case of the H1 vaccine component, the same H1 vaccine strain (A/California/7/2009) was used in each year. These plots were generated using the “geom_point” function from R package “ggplot2” (version 3.4.2). The fold change of HAI titer for each patient was computed by dividing the post-vaccination HAI titer by the corresponding pre-vaccination HAI titer.

### Longitudinal analysis of antibody broadening

To evaluate antibody broadening in response to homologous H1 (A/California/7/2009) we divided the HAI titers for H1N1 viruses from each individual subject into responders and non-responders to each viral strain in each year. Responders were defined by having non-decreasing fold changes of HAI titers (post-vaccination HAI titer / pre-vaccination HAI titer), i.e. fold change of HAI titers greater than 1. The non-responders were defined by having decreasing fold changes of HAI titers (post-vaccination HAI titer / pre-vaccination HAI titer), i.e. fold change of HAI titers less than 1. We tracked patients in the two age groups: >50 years old, having four years of complete HAI titers (2013-2016); and <38 years old, having three years of complete HAI titers (2014-2016). Bar plots showing the responder and non-responder ratios was graphed using “geom_bar” function from R package “ggplot2” (version 3.4.2). Linear regression analyses were also performed on the fraction of responders (or non-responders) in each year after standardizing the responder (or non-responder) value to the number of subjects in each age group (>50 years old or <38 years old). The linear regressions were performed using “ggscatter” function with the fitting equation shown by “stat_regline_equation” function from R package “ggpubr” (version 0.6.0). Corresponding *P* and r^2^ values were computed using “lm” function from R package “stats” (version 4.3.1).

### Immune Reactions in silico

The computational model is adapted from past work on the effects of repeated vaccination with COVID vaccines (Yang *et al*., 2023). Changes to the original model are described in the main text and below, along with key mathematical equations that describe the model. Interested readers can find in-depth rationale for model development, exploration of alternative model structures, and further analysis of parameter sensitivity in the earlier paper on the outcome of multiple immunizations with COVID vaccines (Yang *et al*., 2023). We use the same symbols to denote quantities as in the paper on COVID vaccines (Yang *et al*., 2023).

Differential equations describe antigen dynamics and this is combined with stochastic simulations of GC and EGC processes. The time step is 0.01 day. For each situation, 200 GCs are simulated, and from the second immunization on, 1 EGC is simulated along with the GCs. Ten such simulations are carried out for each set of conditions and the results are averaged over the ten simulations to report results.

### Antigen dynamics

Differential equations describe the reactions that govern the concentration of antigen and antibody, as shown in the table below. We use the following abbreviations and symbols: soluble antigen (Ag), soluble antibody (Ig), soluble immune complex (IC), immune complex on follicular dendritic cell (IC-FDC), plasma cell (PC), rate of decay (d), rate of reaction (k), dissociation constant of serum antibodies (K_d_), dissociation constant of plasma cell BCRs (K^Pc^_d_).

| Equation | Reaction(s) | Description |
| --- | --- | --- |
| $\frac{[Ag][Ig]}{[IC]} = K_d$ | $Ag + Ig \leftrightarrow IC$ | Fast equilibrium for formation of immune complex |
| $\partial_t[Ag] = -d_{Ag}[Ag]$ | $Ag \rightarrow \emptyset$ | Decay of free soluble antigen |
| $\partial_t[IC] = -k_{deposit}[IC]$<br>$\partial_t[IC - FDC] = k_{deposit}[IC]$<br>$- d_{IC}[IC - FDC]$ | $IC \rightarrow IC - FDC$<br>$IC - FDC \rightarrow \emptyset$ | Immune complex transport to follicular dendritic cells<br>Consumption and decay of antigen on follicular dendritic cells |
| $\partial_t[Ig] = k_{Ig}[PC] - d_{Ig}[Ig]$ | $PC \rightarrow PC + Ig$<br>$Ig \rightarrow \emptyset$ | Antibody production by plasma cells<br>Decay of free soluble antigen |
| $\partial_t K_a = \frac{(K_a^{PC} - K_a)k_{Ig}[PC]}{[Ig] + [IC]}$ | - | Derived from equations above, as detailed in supplement of Yang, et al. (2023). |

The parameters used in these equations can be found in Table S2, and are identical to our previous publication (Yang *et al*., 2023). Upon the first vaccination, only weakly binding IgM antibodies are available for binding to soluble antigen and deposoiting ICs on to FDCs. After subsequent vaccinations, antigen specific antibodies are available.

### B cell dynamics in GCs and EGCs

Each GC is associated with a pool of 2000 naïve B cells (Yang *et al*., 2023). A fraction ρ,_i_ of these naïve B cells target epitope ρ,. We model 3 epitopes of the influenza spike protein, which is an increase from the 2 epitopes previously used to model the SARS-CoV-2 spike protein (Yang *et al*., 2023). This allows us to account for the diversity in influenza strains, each with distinct epitopes that may be conserved with the H1N1 CA09 vaccine strain (see main text).

The germline binding affinities *E =, -log(K_d_)* of the naïve B cells for the vaccine strain have discrete values between 6 and 8, expressed as E_k_ = 6 + 0.2k for k = 0,1,…,10. The lower value of 6 was chosen based on data as described in main text, and germline affinities that are cells targeting epitope *i*, in the affinity bin E_k_ is a truncated geometric distribution and is one hundred times greater have also been observed (Yang *et al*., 2023). The frequency of naïve B determined as follows:

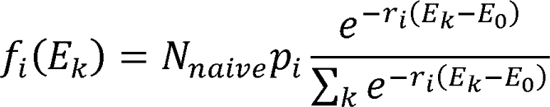

where the number of naïve B cells *N_naive_* is 2000 and the minimum germline affinity E_0_ is 6 in these simulations. r_i_ is determined such that

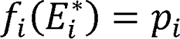

where E^*^_i_ is defined as follows using the parameters E^h^_1_, dE_12_, and dE_13_:

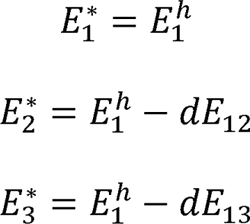

The parameters are set such that E^*^_1_ < E^*^_2_ < E^*^_3_, reflecting the generally higher affinities for more dominant epitopes. The immunodominance hierarchy is further enforced by setting p_1_ < p_2_ < p_3_, meaning that more dominant epitopes are targeted by a greater number of naïve B cells.

Since individuals initially have a weak response (sub-dominant) against historical influenza strains, the germline binding affinity of all naïve B cells that target the epitopes that are conserved between the vaccinating strain and historical strains is set to the lowest possible germline affinity in these simulations (*E* = 6).

After initializing the pool of naïve B cells, the B cells can be stochastically activated. The probability of activation for a naïve B cell depends on the quantity of vaccine antigen it captures, which is determined by both the antigen concentration and binding affinity to the vaccine strain. The amount of antigen captured by B cell *j* is modeled as

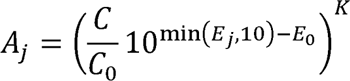

where *C* is the effective antigen concentration, *C*_0_ is the reference antigen concentration, *E*_j_ is the B cell’s binding affinity for vaccine strain, and *E*_0_ is the reference affinity. The effective antigen concentration is *C = 0.01([Ag] + [IC]) + [IC − FDC]*, where antigens presented on FDCs are more potent at activating B cells (Kim et al., 2006). The selection stringency *K* represents how binding affinity. The probability of activation is *P(B cell j is activated) = min(A_j_,1)*.

Activated B cells can then stochastically enter the GC. Entry into the GC depends on antigen captured and competition for limited T cell help (Lee et al., 2021; Schwickert et al., 2011). The rate of entry for an activated B cell *j* is

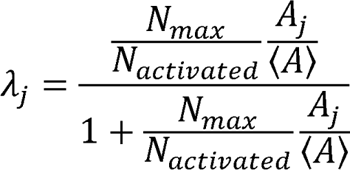

where *N*_activated_ is the number of activated B cells, *N*_max_ is the capacity for GC entry based on limited T cell help, and [A] is the average amount of antigen captured by all B cells. Thus, 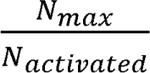 represents the competition between B cells for T cell help and 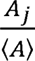 represents the competitive advantage of a particular B cell *j* over other cells. The probability of GC entry is *P(B cell j enters GC) = 1 − e^−yjdt^*.

GC B cells also capture antigen and compete for T cell help to become stochastically activated. The rate of positive selection is

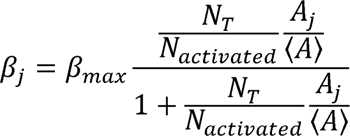

where β_max_ is the maximum rate of positive selection, *N*_activated_ is the number of activated B cells, and *N*_T_ is the number of helper T cells.

If a B cell is positively selected, it exits the GC with probability *p*_exit_ or is recycled for mutation-selection cycles in the GC with probability 1 − *p*_exit_. If the B cell exits, it becomes a plasma cell with probability *p*_plasma_ or a memory cell with probability 1-*p*_plasma_. If the B cell proliferates, one of the daughter cells mutates. The mutation may change affinity (probability 0.2), result in apoptosis (probability 0.3), or be silent (Zhang and Shakhnovich, 2010).

Each B cell has a string of 0s and 1s for the residues on the paratope with a total length of *n*_res_. The string of residues starts as all 0s in a naïve B cell. When there is an affinity-changing mutation, one of the bits (residues) is randomly chosen and flipped. The change in affinity is Gromiha, 2006; Zhang and Shakhnovich, 2010). The affinity of a B cell *j* for a particular strain is drawn from a shifted log-normal distribution, independently for each residue (Kumar and determined by both the germline affinity and affinity-changing mutations, as follows:

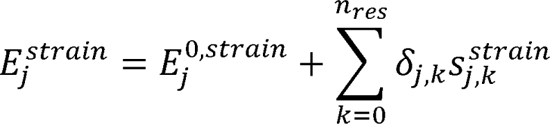

where *E*^o,strain^_j_ is the germline affinity, δ_j,k_ ∈ {0,1} is the mutational state of residue *k*, and *S*^strain^_j,k_ is the change in affinity due to a mutation at residue *k*. *S*^strain^_j,k_ is correlated between different strains, as follows (*S*^1^_j,k_ is the change in affinity for strain 1, *S*^2^_j,k_ is the change in affinity for strain 2, and so on). The values of *S*^strain^_j,k_ are drawn from identical log normal distributions that are correlated as follows:

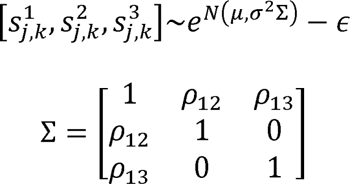

where μ,σ,∈ were chosen such that only ∼5 % of affinity-changing mutations are beneficial, as shown in experimental studies (Kumar and Gromiha, 2006; Zhang and Shakhnovich, 2010). Σ is the correlation matrix and ρ_12_ and ρ_13_ parameterize the correlation of affinity changes between conservation between the strains for B cell *j*’s target epitope, as described in the main text. We strains 1 and 2 and strains 1 and 3, respectively. The level of correlation is related to the level of only consider correlation between the vaccine strain (strain 1) and historical strains (strains 2 and 3), but not between strains 2 and 3. Since B cells are selected for their affinity to strain 1, the correlation between strains 2 and 3 does not impact the nature of the antibody or memory B cell response.

In this study, we considered the correlation between three strains compared to two strains in our COVID model (Yang *et al*., 2023) since we examined the effect of sequential immunization on B cell responses against multiple unmatched influenza strains.

After the first immunization, pre-existing memory cells stochastically expand and differentiate in the EGC. The memory cells are selected for expansion in the same affinity-dependent way as GC B cells, except memory cells do not undergo mutation in our model. The number of helper T cells is set to its maximum value to reflect the faster kinetics of the EGC (Goel *et al*., 2021; Moran et al., 2019). B cells exiting the EGC differentiate into plasma cells with a probability of 0.6 (Moran *et al*., 2018).

### Epitope masking

When epitope masking is considered, GC B cells specific for a particular epitope cannot capture antigen on FDCs if that epitope is bound to circulating antibodies. These circulating antibodies are produced by plasma cells from previous immunizations and expansion and differentiation of memory cells from previous immunizations. The amount of bound antigen is calculated using fast equilibrium of receptor-ligand binding:

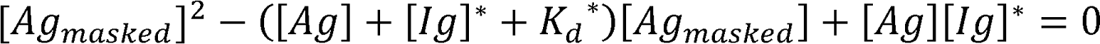

where [Ag_masked_], is the concentration of masked, i.e. bound, antigen, [Ig]* is the effective antibody concentration, and K_d_* is the effective average dissociation constant. The values in the overlap, [Ig]* and K_d_* are exactly the concentration of antibodies targeting a particular epitope equation above are calculated separately for each epitope. When we do not consider epitope [Ig]* and K_d_* the average dissociation constant of those antibodies. In the presence of epitope overlap, some antibodies can mask epitopes that spatially overlap with their primary target. In this case

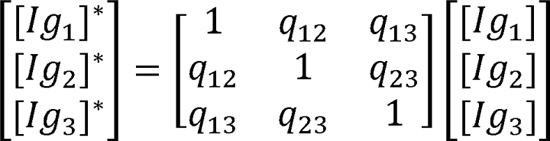

where [Ig_i_]* is the effective concentration of antibodies targeting epitope *i* and [Ig_1_], is the actual concentration of antibodies targeting epitope *i.q_mm_*. In describes the overlap between epitope *m* and *n*is the fraction of antibodies targeting epitope *m* that can mask epitope *n* (and vice versa). The effective average dissociation constant *K_d,i_*\* is calculated similarly:

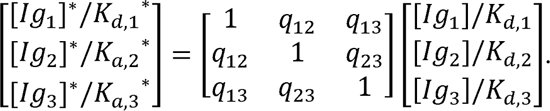

The concentration of bound antigen [Ag_masked_], is then calculated and subtracted from the total antigen concentration since bound antigen cannot be seen by B cells. The resulting antigen concentration is then scaled such that the fraction of soluble antigen and fraction of antigen on the FDC match those fractions before epitope masking is not considered.

### Modifications to the original model

As a summary, the revised model includes three epitopes instead of two epitopes and considers three strains instead of two. This manifests in the distribution of naïve B cells’ germline affinities, the effective antigen concentration in epitope masking, and the correlated affinity changes in the GC. Using multiple epitopes and strains accounts for the diversity in influenza strains, each with distinct epitopes that may be conserved with the vaccine strain. By examining the relationships between multiple epitopes and strains, we gained general insights into factors that affect the number of immunizations needed to achieve a given level of coverage for historical and emergent strains, as detailed in the main text.

## Notes

### Funding Statement

The authors acknowledge support form the NIH [U19AI057229; AI155447, AI137057, AI153098, AI146779, AI089618, AI060354; contracts 75N93019C00050 and 75N93019C00052.

### Author Declarations

Nunez, I.A., Carlock, M.A., Allen, J.D., Owino, S.O., Moehling, K.K., Nowalk, P., Susick, M., Diagle, K., Sweeney, K., Mundle, S., et al. (2017). Impact of age and pre-existing influenza immune responses in humans receiving split inactivated influenza vaccine on the induction of the breadth of antibodies to influenza A strains. PloS one 12, e0185666. 10.1371/journal.pone.0185666. In this 2017 study, the study procedures, informed consent, and data collection documents were reviewed and approved by was approved by the Western Institutional Review Board and the Institutional Review Boards of the University of Pittsburgh and the University of Georgia. Informed written consent was obtained from the parents/guardians of the children

