## Supplementary figures and images for "Repeated vaccination with homologous influenza hemagglutinin broadens human antibody responses to unmatched flu viruses"

### Figures S1 and S2

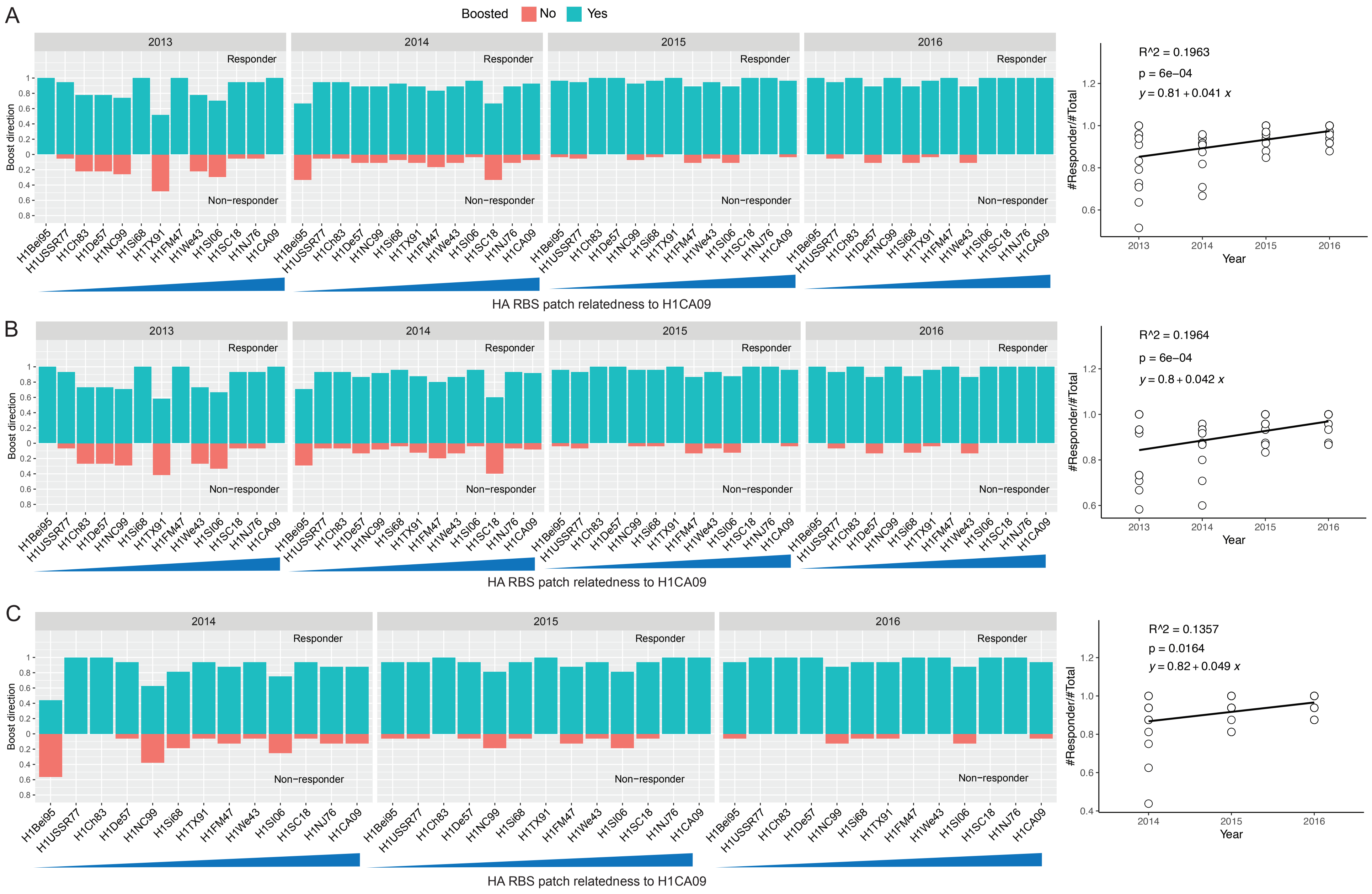

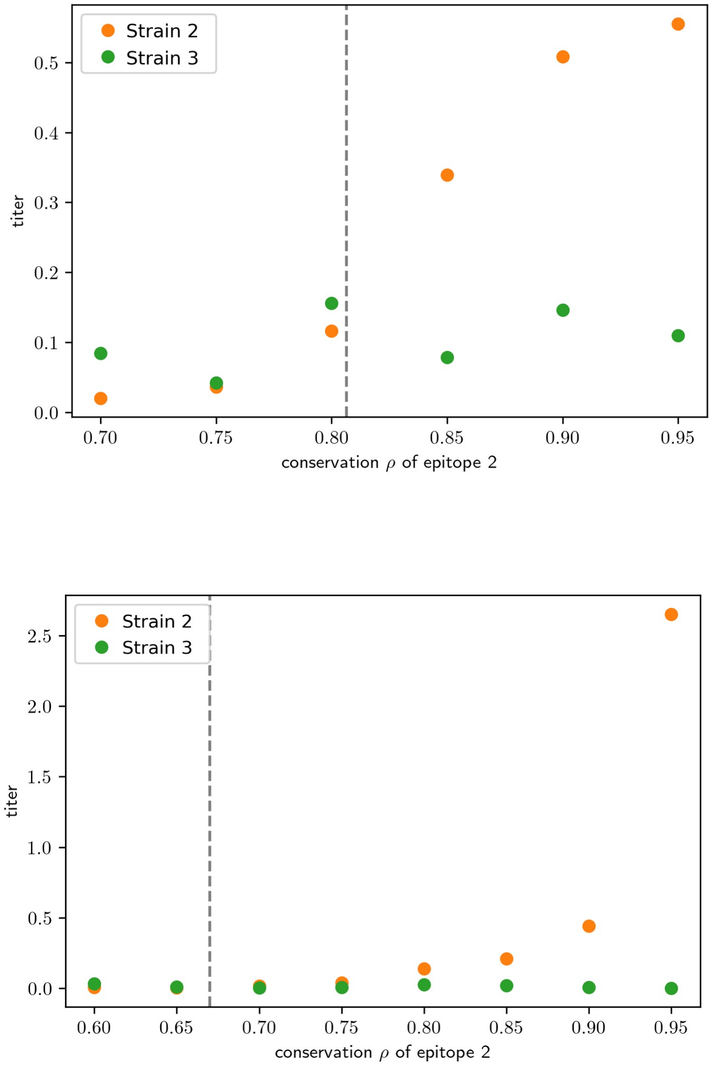
