## Supplementary material for "Repeated vaccination with homologous influenza hemagglutinin broadens human antibody responses to unmatched flu viruses": Tables S1 and S2

Table S1: HA sequence information for the influenza stains in this study

| **Strain** | **Genbank** | **Type** | **Year** | **Location** | **GISAID** |
| --- | --- | --- | --- | --- | --- |
| H1N1 A_Beijing_262_1995 | AB304819.1 | H1N1 | 1995 | Beijing |  |
| H1N1 A_Brazil_11_1978 | HQ008267.1 | H1N1 | 1978 | Brazil |  |
| H1N1 A_Brisbane_59_2007 | JN899402.1 | H1N1 | 2007 | Brisbane |  |
| H3N2 A_Brisbane_10_2007 | KM978061.1 | H3N2 | 2007 | Brisbane |  |
| B_Brisbane_60_2008 | FJ766842.1 | B | 2008 | Brisbane |  |
| H1N1 A_California_07_2009 | NC_026433.1 | H1N1 | 2009 | California |  |
| H1N1 A_California_10_1978 | CY021717.1 | H1N1 | 1978 | California |  |
| H1N1 A_Chile_1_1983 | CY121261.1 | H1N1 | 1983 | Chile |  |
| B_Colorado_06_2017 | CY236607.1 | B | 2017 | Colorado |  |
| H1N1 A_Denver_1957 | CY146793.1 | H1N1 | 1957 | Denver |  |
| B_Florida_4_2006 | EU515992.1 | B | 2006 | Florida |  |
| H1N1 A_Fort_Monmouth_1_1947 | AF494250.1 | H1N1 | 1947 | Fort_Monmouth |  |
| H3N2 A_Fujian_411_2002 | EU501153.1 | H3N2 | 2002 | Fujian |  |
| B_Harbin_7_1994 | CY040441.1 | B | 1994 | Harbin |  |
| B_Hong_Kong_330_2001 | AF532549.1 | B | 2001 | Hong_Kong |  |
| H3N2 A_Hong_Kong_1_1968 | AF348177.1 | H3N2 | 1968 | Hong_Kong |  |
| H3N2 A_Hong_Kong_4801_2014 |  | H3N2 | 2014 | Hong_Kong | EPI1026711 |
| H3N2 A_Kentucky_UR07-0028_2008 | CY037791.1 | H3N2 | 2008 | Kentucky_UR07-0028 |  |
| B_Lee_1940 | K00423.1 | B | 1940 | Lee |  |
| B_Malaysia_2506_2004 | EU124275.1 | B | 2004 | Malaysia |  |
| B_Massachusetts_2_2012 | MT056027.1 | B | 2012 | Massachusetts |  |
| H1N1 A_Michigan_45_2015 | KY117023.1 | H1N1 | 2015 | Michigan |  |
| H3N2 A_Mississippi_1_1985 | L19003.1 | H3N2 | 1985 | Mississippi |  |
| H3N2 A_Nanchang_933_1995 | CY108293.1 | H3N2 | 1995 | Nanchang |  |
| H1N1 A_New_Caledonia_29_1999 | DQ508857.1 | H1N1 | 1999 | New_Caledonia |  |
| H1N1 A_New_Jersey_1976 | CY147422.1 | H1N1 | 1976 | New_Jersey |  |
| H3N2 A_New_York_55_2004 | KM821338.1 | H3N2 | 2004 | New_York |  |
| H3N2 A_Panama_2007_1999 | EF626612.1 | H3N2 | 1999 | Panama |  |
| H3N2 A_Perth_16_2009 | GQ293081.1 | H3N2 | 2009 | Perth |  |
| B_Phuket_3073_2013 |  | B | 2013 | Phuket | EPI2195537 |
| H3N2 A_Port_Chalmers_12_1973 | CY113109.1 | H3N2 | 1973 | Port_Chalmers |  |
| H1N1 A_Puerto_Rico_8_1934 | EF467821.1 | H1N1 | 1934 | Puerto_Rico |  |
| H3N2 A_Shangdong_9_1993 | Z46417.1 | H3N2 | 1993 | Shangdong |  |
| B_Sichuan_379_1999 | EF566113.1 | B | 1999 | Sichuan |  |
| H3N2 A_Sichuan_30_1989 | CY108211.1 | H3N2 | 1989 | Sichuan |  |
| H1N1 A_Singapore_6_1986 | D00406.1 | H1N1 | 1986 | Singapore |  |
| H1N1 A_Solomon_Islands_03_2006 | EU100724.1 | H1N1 | 2006 | Solomon_Islands |  |
| H1N1 A_South_Carolina_1_1918 | AF117241.1 | H1N1 | 1918 | South_Carolina |  |
| H3N2 A_Switzerland_9715293_2013 |  | H3N2 | 2013 | Switzerland | EPI814528 |
| H3N2 A_Sydney_5_1997 | KM821316.1 | H3N2 | 1997 | Sydney |  |
| H1N1 A_Texas_36_1991 | DQ508889.1 | H1N1 | 1991 | Texas |  |
| H3N2 A_Texas_1_1977 | EF626623.1 | H3N2 | 1977 | Texas |  |
| H3N2 A_Texas_50_2012 | KC892952.1 | H3N2 | 2012 | Texas |  |
| B_Texas_06_2011 | KC813979.1 | B | 2011 | Texas |  |
| H1N1 A_USSR_90_1977 | HQ008265.1 | H1N1 | 1977 | USSR |  |
| H3N2 A_Victoria_361_2011 | KM821347.1 | H3N2 | 2011 | Victoria |  |
| H1N1 A_Weiss_1943 | CY147366.1 | H1N1 | 1943 | Weiss |  |
| H3N2 A_Wisconsin_67_2005 | CY163704.1 | H3N2 | 2005 | Wisconsin |  |
| B_Wisconsin_1_2010 | CY115183.1 | B | 2010 | Wisconsin |  |
| B_Yamagata_16_1988 | M36105.1 | B | 1988 | Yamagata |  |

**Table S2:** Simulation parameters. Highlighted parameters were modified from the original model.

| **­Parameter** | **Value** | **Description** |
| --- | --- | --- |
| **Antigen and antibody dynamics** | | |
| $k_{Ig}$ | $0.8 \times{10}^{-2}$ nM day^-1^ PC^-1^ | Rate of antibody production per plasma cell per day |
| $d_{Ig}$ | 0.025 day^-1^ | Antibody decay rate |
| $d_{Ag}$ | 3 day^-1^ | Antigen decay rate |
| $k_{deposit}$ | 1 hour^-1^ | Rate of immune complex transport to FDC |
| $d_{IC}$ | 0.15 day^-1^ | Rate of decay of immune complex on FDC |
| ${[Ag]}_{0}$ | 0 nM | Initial Conditions |
| ${[Ig]}_{0}$ | 10^-2^ nM |  |
| ${[IC]}_{0}$  ${[IC-FDC]}_{0}$ | 0 nM |  |
| **B cell affinities** | | |
| $N_{naive}$ | 2000 cells/GC | Number of naïve B cells per GC |
| $p_{1}$ | 0.8 | Fraction of naïve B cells that target epitope 1 |
| $p_{2}$ | varied: 0.15, 0.18 | Fraction of naïve B cells that target epitope 2 |
| $p_{3}$ | varied: 0.05, 0.02 | Fraction of naïve B cells that target epitope 3 |
| $E_{1}^{h}$ | 7 | Parameter for the germline affinity distribution of epitope 1. Affinity at which there is on average one naïve B cell targeting epitope 1 available for each GC |
| ${dE}_{12}$ | 0.4 | Parameter for the germline affinity distribution of epitope 2. $E_{1}^{h}- {dE}_{12}$ is the affinity at which there is on average one naïve B cell targeting epitope 2 available for each GC |
| ${dE}_{13}$ | 0.6 | Parameter for the germline affinity distribution of epitope 3. $E_{1}^{h}- {dE}_{13}$ is the affinity at which there is on average one naïve B cell targeting epitope 2 available for each GC |
| $n_{res}$ | 80 | Length of string representation of B cell residues, upper range of sum of CDR lengths in light and heavy chain (Nowak et al. 2016) |
| $\mu$, $\sigma$, $\epsilon$ | 3.1, 1.2, 3.08 | Parameters for shifted log-normal distribution that models the effect of affinity-changing mutations |
| $\rho_{12}$ | epitope 1: 0.4  epitope 2: typically 0.95, varied: 0.6-0.95 in Fig. 8  epitope 3: 0.4 | Level of conservation between strain 1 and 2 |
| $\rho_{13}$ | epitope 1: 0.4  epitope 2: 0.4  epitope 3: 0.95 | Level of conservation between strain 1 and 3 |
| $q_{12}$ | 0 without overlap  0.3 with overlap | Fraction of antibodies that target epitope 1 that can mask epitope 2 and vice versa (i.e. the amount of spatial overlap between epitope 1 and 2) |
| $q_{13}$ | 0 without overlap  0.3 with overlap | Fraction of antibodies that target epitope 1 that can mask epitope 3 and vice versa (i.e. the amount of spatial overlap between epitope 1 and 3) |
| $q_{23}$ | 0 | Fraction of antibodies that target epitope 2 that can mask epitope 3 and vice versa (i.e. the amount of spatial overlap between epitope 2 and 3) |
| **GC and EGC dynamics** | | |
| $C_{0}$ | $8\times{10}^{-3}$ nM | Reference antigen concentration |
| $E_{0}$ | 6 | Reference binding affinity |
| $K$ | 0.5 | Stringency of selection of naïve and GC B cells by helper T cells based on amount of captured antigen |
| $N_{max}$ | 10 day^-1^ | Approximately the maximum number of naïve B cells that can enter the GC per day |
| $N_{T0}$ | 1200 | Maximum number of helper T cells involved in positive selection in GC and EGC |
| $\beta_{max}$ | 2.5 day^-1^ | Maximum rate of positive selection for GC and EGC B cells |
| $\alpha$ | 0.5 day^-1^ | Death rate of GC B cells |
| **Plasma and memory cell dynamics** | | |
| $p_{exit}$ | 0.05 | Probability that a positively selected GC B cell exits and differentiates |
| $p_{plasma}$ | 0.1 | Probability that a differentiating GC B cell becomes a plasma cell |
| $p_{plasma}^{EGC}$ | 0.6 | Probability that a proliferating memory cell in EGC differentiates into a plasma cell |
| $d_{PC}$ | 0.17 day^-1^ | Death rate of plasma cells |
