## Supplemental Information: Data S1 information; Figure S1 and S2 legends for "Repeated vaccination with homologous influenza hemagglutinin broadens human antibody responses to unmatched flu viruses"

**Data S1.**  HAI values for the influenza virus strains, measured across longitudinal vaccine study (2013-2016) for patients >50 years of age and <38 years of age.

**Figure S1. H1N1** **responders regressed over the vaccine.** Responders (green) versus non-responders (red) within each year is graphed for each H1N1 strain in the HAI panel, as in Figure 3. (A) Yearly response for all longitudinally analyzed individuals; at right is a linear regression of the proportion of non-responders against over the four year vaccine data (*P*=6e-04). (B) Data for subjects >50 years in age (*P*=6e-04, linear regression). (C) Data for subjects <38 years in age (*P=*0.0164, linear regression).

**Figure S2. The interplay between conservation of sub-dominant epitopes between historical strains and the immunizing strain and germline frequencies of B cells that target these epitopes**. We vary the conservation $\rho$ of epitope 2 between the immunizing strain and strain 2 while fixing the fraction of naive B cells $p_{i}$*.* If epitope 2 is significantly less conserved between strains 1 and 2 than epitope 3 is between strains 1 and 3, the effects of weaker conservation outweigh the effects of a more favorable germline distribution, resulting in worse titers against strain 2 than against strain 3. The point at which the advantage of the immunodominance hierarchy is overcome by the effects of lower conservation depends on the germline B cell properties. The effects of epitope masking are not considered here. (A) Here, we fix the fraction of naive B cells $p_{i}$ targeting epitope i to $p_{1}=0.8, p_{2}=0.18, p_{3}=0.05$. The conservation $\rho$ of epitope 2 between the immunizing strain and strain 2 is varied, and the titers after the second vaccination are shown. When $\rho$ is greater than the crossover point ~0.8, there are higher titers against strain 2 than strain 3. This is the same order as the order of the corresponding conserved epitope in the immunodominance hierarchy; in particular, epitope 2 (which is conserved with strain 2) is more immunodominant than epitope 3 (which is conserved with strain 3). When $\rho$ is less than the crossover point ~0.8, there are higher titers against strain 3 than strain 2, which is not the same order as the immunodominance hierarchy for the corresponding epitopes. (B) When we alter the relative dominance of the epitopes by changing the fraction of naive B cells targeting each epitope to $p_{1}=0.8, p_{2}=0.18, p_{3}=0.02$, the crossover point occurs at a lower conservation value $\rho\approx0.66$.
